# *Plasmodium falciparum* malaria during pregnancy: the impact of parasitaemia and anaemia on birthweight

**DOI:** 10.1101/2021.08.12.21261986

**Authors:** Dawood Ackom Abbas, Abdul-Hakim Mutala, Samuel Kekeli Agordzo, Christian Kwasi Owusu, Bernard Walter Lartekwei Lawson, Kingsley Badu

**Affiliations:** Department of Theoretical and Applied Biology, Kwame Nkrumah University of Science and Technology, Kumasi, Ghana; Kumasi Center for Collaborative Research for Tropical Medicine, Kwame Nkrumah University of Science and Technology, Kumasi, Ashanti, Ghana

**Keywords:** pregnant women, malaria, anaemia, low birthweight, Sulphadoxine-pyrimethamine, Ghana

## Abstract

Malaria in pregnancy remains a major problem of public health concern in Sub-Saharan Africa due to its endemicity and the diverse consequences on both the mother and the baby. Much attention, therefore, is needed to fully understand the epidemiology of the disease and to mitigate the devastating outcomes. The present study aimed at investigating malaria in pregnancy, its adverse effects on pregnant women and the impact on birthweight of babies. A total of 222 pregnant women gave their consent and were recruited into the study during their routine Antenatal care visits. This study employed a combination of cross-sectional and longitudinal cohort study designs. For 122 women in the cross-sectional arm, blood samples and data were obtained once, whilst 100 women in the longitudinal cohort arm were followed up from recruitment until delivery. Demographic information, obstetric history and risk factors were obtained by administering questionnaires. About 1.0 ml of venous blood was drawn to determine malaria parasitaemia and anaemia status of the participants. The birthweights of the babies were also taken at delivery. The prevalence of malaria and anaemia was 19.8% and 27.0% respectively at registration for all 222 participants. All infections were *P. falciparum* malaria. One hundred and forty-six (65.8%) of participants had ITN but only 72 (32.4%) used it the previous night. Young age and rural settings were risk factors for malaria. Young age and malaria positive pregnant women had increased risk of anaemia. In the follow-up group which ended with 54 participants, the overall prevalence of malaria and anaemia were 18.7% and 32.4% respectively. Fifty-two (96.3%) of pregnant women attended ANC ≥ 4 times and 55.6% took ≥ 3 doses of SP. There were two cases of miscarriage. Low birthweight occurred in 5.6% of babies. Both malaria and anaemia during pregnancy had no significant impact on birthweight of the babies. Although few of the babies had low birthweight, this number can be further reduced when pregnant women attend ANC and take SP at the recommended number of times.

## 1. Introduction

About 90% of the global malaria cases occur in Sub-Saharan Africa including Ghana, with an estimated 25 million women predisposed to the risks posed by the infection and its accompanying adverse consequences during pregnancy (Ibrahim et al., 2017). In Ghana, there is an estimated 3.2 million malaria cases leading to about 38,000 deaths every year (Owusu Adjah and Panayiotou, 2014). It accounts for more than 44% of outpatient department attendance of which an estimated 13.8% are represented by pregnant women resulting in 9.4% of all maternal deaths (Orish et al., 2015; Owusu-Agyei et al., 2007; Saaka et al., 2009). Malaria is also a major contributor to the neonatal mortality rate in Ghana, standing at 29 per 1,000 deaths (Ghana Health Service, 2021).

In view of this, the World Health Organization (WHO) promotes a multi-pronged approach to malaria control during pregnancy. This involves the provision of intermittent preventive treatment with sulphadoxine-pyrimethamine (IPT-SP), use of insecticide treated nets (ITN) and effective case management of confirmed infections (WHO, 2014).

These strategies coupled with other efforts should see a huge reduction in the burden of the infection, however, this is not the case in Ghana. The prevalence of anaemia and malaria, especially asymptomatic malaria, and their deleterious effects during pregnancy continue to be of public health concern in the country. Even though antenatal care attendance in many health facilities in Ghana have seen an increase with the implementation of these strategies, the maternal mortality rate is approximately 308 per 100,000 live births (WHO et al., 2019).

A study conducted in the Ashanti region found a prevalence of 12.6% and 62.6% of malaria and anaemia respectively in pregnant women (Tay et al., 2013). A previous study also recorded a 15% prevalence of the infection among pregnant women in the same region (Tutu et al., 2011a). Other studies also reported more anaemia cases in urban settings compared to rural settings (Iqbal et al., 2016). These results emphasize the need to investigate the underlying cause of the infection amidst the several control measures. Studies conducted in the region are mostly cross-sectional, examining malaria and its associated anaemia among pregnant women. A longitudinal study is therefore needed to investigate the intensity of the infection and its consequence during the gestation period and the ultimate outcome.

The present study thus sought to investigate the burden of malaria during pregnancy, its adverse effects on the pregnant women and the impact on birth outcome and some factors that influence the infection prevention and control.

## 2. Methods

### 2.1 Study areas

The study was carried out in four health facilities namely: the University (KNUST) Hospital, Aniniwah Medical Centre (Emena), Kuntanase Government Hospital and Agona Government Hospital, all in the Ashanti region of Ghana from February to December 2018.

The University (KNUST) Hospital and Aniniwah Medical Centre (Emena) are located in Kumasi in the Oforikrom Municipality. Kuntanase Government Hospital is located in Kuntanase which is the capital of the Bosomtwe district; the Agona Government Hospital is located in Agona, the capital of Sekyere-South district. Both Kuntanase Government Hospital and Agona Government Hospital serve as the health care centres to their respective towns and the nearby towns and villages within their districts.

### 2.2 Ethical consideration

Prior to the commencement of this study, ethical approval was obtained from the Committee on Human Research Publications and Ethics (CHRPE) of the School of Medical Sciences, Kwame Nkrumah University of Science and Technology (KNUST), and Komfo Anokye Teaching Hospital (Ref: CHRPE/AP/544/18). All the study participants signed informed consent forms after the study goals were thoroughly explained to them in the local language, where required.

### 2.3 Study design and patients

The study population were pregnant women seeking antenatal care either for the first time or were present for their routine monthly visits. Two hundred and twenty-two (222) participants were recruited at enrolment out of which 100 of them were selected for follow up till delivery. Inclusion criteria were: informed consent, no critical illness and additionally, delivery at the hospital for the follow-up group. Demographic information, the use of preventive measures and obstetric history were taken through questionnaire administration. The malaria and anaemia status of all participants recruited were determined. For the follow-up group, malaria and anaemia status were determined for a maximum of 3 times till delivery to coincide with their scheduled visits at the ANC centre. Birthweight record of babies was taken at delivery.

### 2.4 Sample collection and examination

About 1.0 ml of venous blood samples were taken by a trained phlebotomist using sterile syringes and needles and transferred into well labelled EDTA tubes. Each blood sample was screened for malaria parasites using RDT and microscopy, and their haemoglobin concentration (anaemia status) was determined using Sysmex XP-300.

Thick smears of the blood samples were prepared, stained with 10% Giemsa solution for 15 minutes and examined under the microscope (x100 objective lens) for the presence of malaria parasites. The parasites were quantified by counting parasites against white blood cells (WBC). Depending on the number of parasites present, the parasites were counted against 200 or 500 white blood cells. A blood film is declared negative when no parasite was seen after 100 high power fields have been viewed (WHO, 2010).

Full blood counts were done to estimate the haemoglobin concentration of the blood samples using the Sysmex XP-300 automated haematology analyser. Concentrations lesser than 11 g/dl were reported as anaemic (WHO, 2001).

Data on ANC visits, SP taken, and birth weights of babies were taken from the delivery records for women who were involved in the follow up. Birth weights <2.5 kg were considered low and weights > 2.5 kg but less than 4.0 kg were considered normal (WHO, 2016).

### 2.5 Data analysis

Data collected were coded and entered into Microsoft Office Excel 2016. The data was then analysed using Microsoft Office Excel 2016 and SPSS version 23 software packages.

Tables showing percentages were provided for categorical variables and compared using the Pearson’s Chi-square test. Means and standard deviations were provided for continuous variables. Binary logistic regression was used to analyse the strength of association of dependent variables with malaria, anaemia, and low birthweight. Mann Whitney U test and Kruskal Wallis tests were used to compare non-parametric variables where applicable. For all tests, p values less than 0.05 were considered statistically significant.

## 3.0 Results

### 3.1 Maternal characteristics of participants

Overall, 222 pregnant women were recruited for the study with a mean age of 28.2 years. As shown in Table 1 below, majority of the participants were above 20 years old and multigravidae. Most of them were in their 2^nd^ trimester, possessed ITN and had not taken IPT-SP.

**Table 1.**
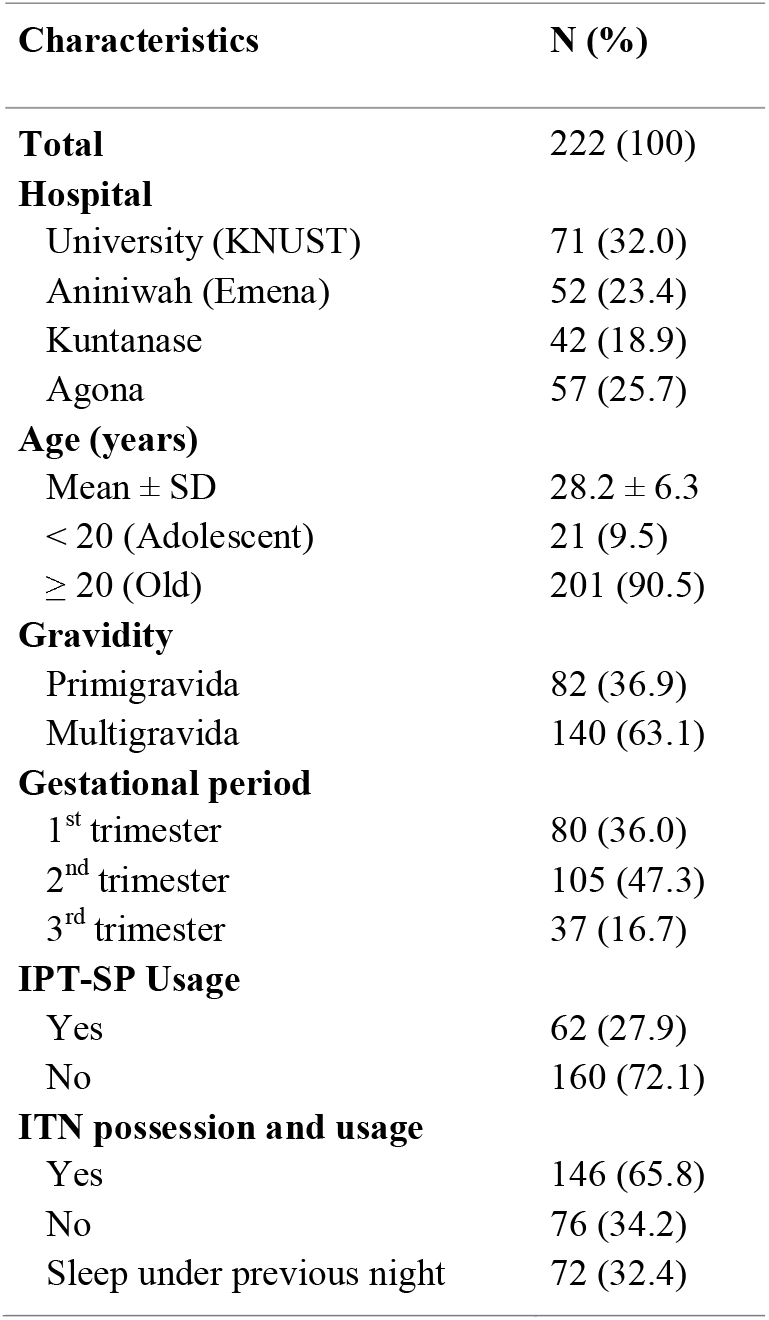
Maternal characteristics of participants recruited for the study

### 3.2 Prevalence of malaria and anaemia

The overall prevalence of malaria and anaemia at enrolment were 19.8% and 27.0% respectively. All recorded malaria infections were caused by *P. falciparum*.

#### 3.2.1 Hospital based prevalence

The highest malaria prevalence of 26.3% was recorded at the Agona Government hospital whereas Aniniwah Medical Centre (Emena) recorded the highest for anaemia prevalence of 40.4%. The least malaria and anaemia prevalence of 7.0% and 9.9% respectively were recorded at the University hospital (Fig 1). Pregnant women from Kuntanase and Agona government hospitals were found to be at higher risk of infection (Table 3). Additionally, participants with malaria had increased risk of being anaemic (OR=4.733 95% CI 2.357-9.504, p<0.001) (Table 3).

**Table 2:**
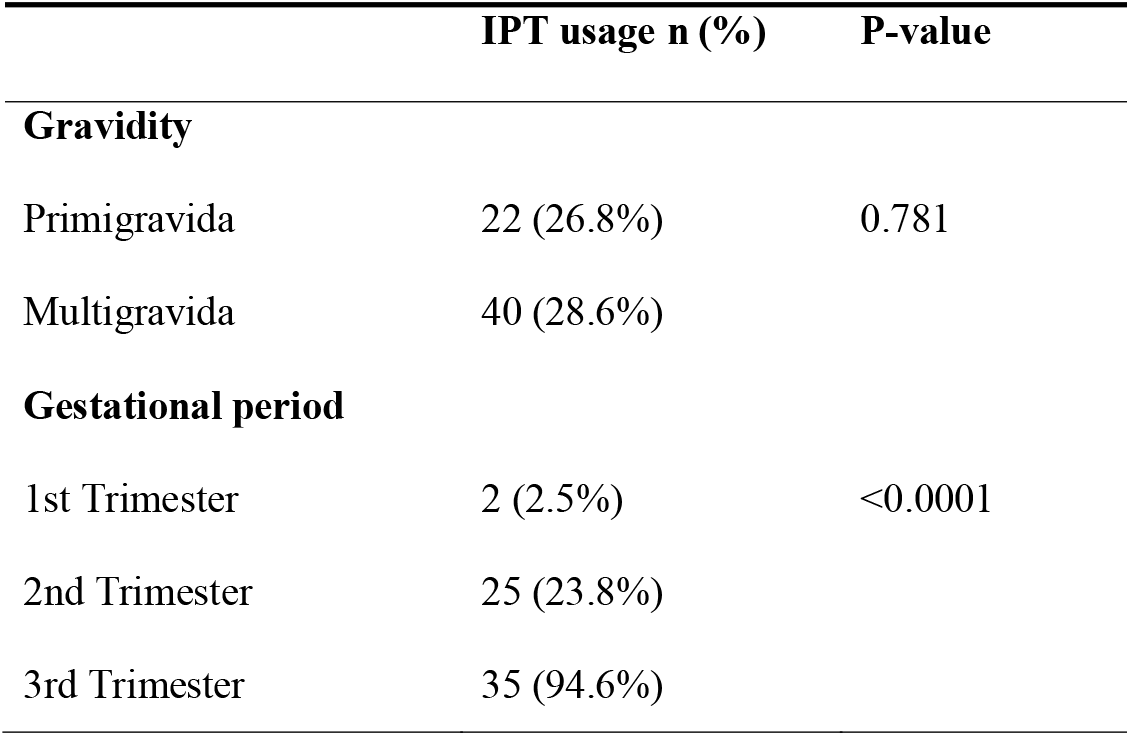
IPT usage among participants.

**Table 3:**
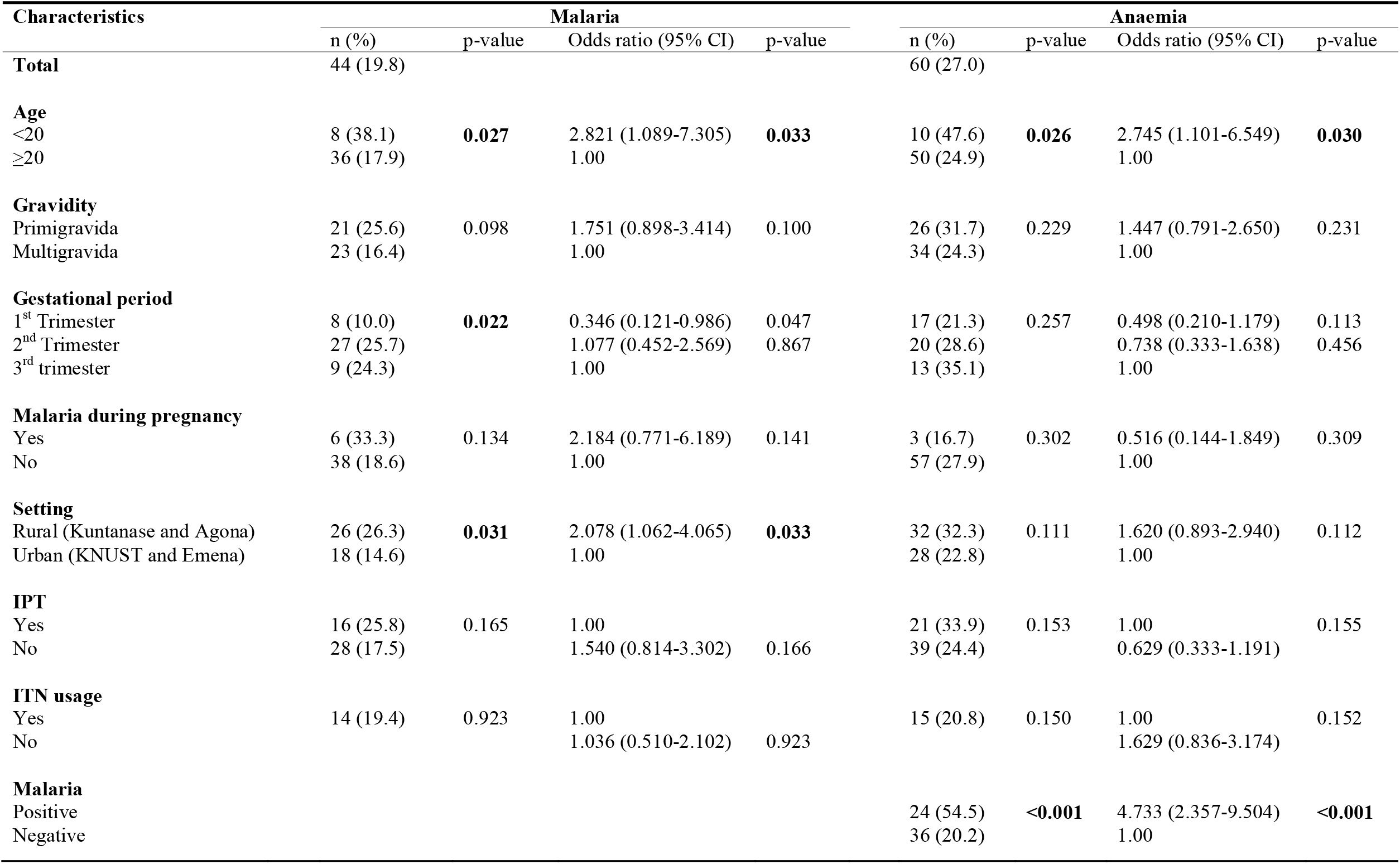
Maternal characteristics and binary logistic regression analysis on factors associated with malaria and anaemia

| Characteristics | Malaria |  |  |  | Anaemia |  |  |  |
| --- | --- | --- | --- | --- | --- | --- | --- | --- |
|  | n (%) | p-value | Odds ratio (95% CI) | p-value | n (%) | p-value | Odds ratio (95% CI) | p-value |
| **Total** | 44 (19.8) |  |  |  | 60 (27.0) |  |  |  |
| **Age** |  |  |  |  |  |  |  |  |
| <20 | 8 (38.1) | **0.027** | 2.821 (1.089-7.305) | **0.033** | 10 (47.6) | **0.026** | 2.745 (1.101-6.549) | **0.030** |
| ≥20 | 36 (17.9) |  | 1.00 |  | 50 (24.9) |  | 1.00 |  |
| **Gravidity** |  |  |  |  |  |  |  |  |
| Primigravida | 21 (25.6) | 0.098 | 1.751 (0.898-3.414) | 0.100 | 26 (31.7) | 0.229 | 1.447 (0.791-2.650) | 0.231 |
| Multigravida | 23 (16.4) |  | 1.00 |  | 34 (24.3) |  | 1.00 |  |
| **Gestational period** |  |  |  |  |  |  |  |  |
| 1<sup>st</sup> Trimester | 8 (10.0) | **0.022** | 0.346 (0.121-0.986) | 0.047 | 17 (21.3) | 0.257 | 0.498 (0.210-1.179) | 0.113 |
| 2<sup>nd</sup> Trimester | 27 (25.7) |  | 1.077 (0.452-2.569) | 0.867 | 20 (28.6) |  | 0.738 (0.333-1.638) | 0.456 |
| 3<sup>rd</sup> trimester | 9 (24.3) |  | 1.00 |  | 13 (35.1) |  | 1.00 |  |
| **Malaria during pregnancy** |  |  |  |  |  |  |  |  |
| Yes | 6 (33.3) | 0.134 | 2.184 (0.771-6.189) | 0.141 | 3 (16.7) | 0.302 | 0.516 (0.144-1.849) | 0.309 |
| No | 38 (18.6) |  | 1.00 |  | 57 (27.9) |  | 1.00 |  |
| **Setting** |  |  |  |  |  |  |  |  |
| Rural (Kuntanase and Agona) | 26 (26.3) | **0.031** | 2.078 (1.062-4.065) | **0.033** | 32 (32.3) | 0.111 | 1.620 (0.893-2.940) | 0.112 |
| Urban (KNUST and Emena) | 18 (14.6) |  | 1.00 |  | 28 (22.8) |  | 1.00 |  |
| **IPT** |  |  |  |  |  |  |  |  |
| Yes | 16 (25.8) | 0.165 | 1.00 |  | 21 (33.9) | 0.153 | 1.00 | 0.155 |
| No | 28 (17.5) |  | 1.540 (0.814-3.302) | 0.166 | 39 (24.4) |  | 0.629 (0.333-1.191) |  |
| **ITN usage** |  |  |  |  |  |  |  |  |
| Yes | 14 (19.4) | 0.923 | 1.00 |  | 15 (20.8) | 0.150 | 1.00 | 0.152 |
| No |  |  | 1.036 (0.510-2.102) | 0.923 |  |  | 1.629 (0.836-3.174) |  |
| **Malaria** |  |  |  |  |  |  |  |  |
| Positive |  |  |  |  | 24 (54.5) | **<0.001** | 4.733 (2.357-9.504) | **<0.001** |
| Negative |  |  |  |  | 36 (20.2) |  | 1.00 |  |

**Fig 1.**
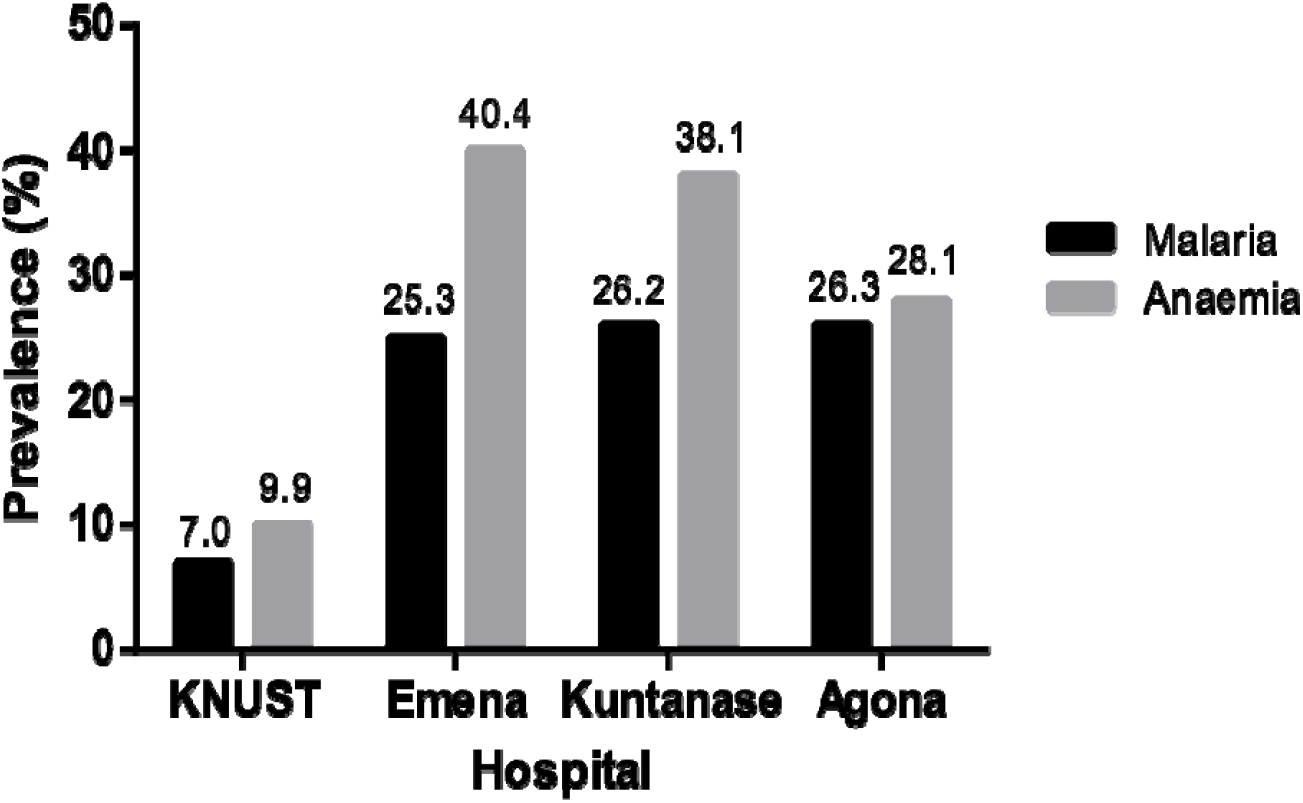
Prevalence of malaria and anaemia in the various hospitals.

#### 3.2.2 Age prevalence

The age range of all the participants was between 15 and 45 years with a mean age of 28.2 ± 6.3 years. This was further categorized into two; young (adolescent) and old, i.e. below twenty (20) years of age, and twenty and above (≥20) respectively. Among the age groups, malaria prevalence was higher (38.1%) in those aged <20 compared to 17.9% in pregnant women aged ≥20. Anaemia was also more prevalent amongst young pregnant women than in the older women, 10 (47.6%) and 50 (24.9%) respectively. Pregnant women aged less than 20 had increased odds of both malaria and anaemia (Table 3).

#### 3.2.3 Gravidity and gestational prevalence

Primigravida women were eighty-two (36.9%) whilst multigravida women were one hundred and forty (63.1%). Twenty-one (25.6%) of primigravida women and twenty-three (16.4%) of multigravida women had malaria parasitaemia. Anaemia prevalence was 31.7% in primigravida women and 24.3% in multigravida women (Table 3).

Based on their gestational period, 80 (36.0%) women were in their first trimester, 105 (47.3%) in the second trimester and 37 (16.7%) in their third trimester. More than half (61.4%) of the participants who tested positive for malaria were in their second trimester with the least recorded in women who were in their first trimester (18.2%). Anaemia prevalence was 35.1% in third trimester, 28.6% in second trimester and 21.3% in first trimester (Table 3).

#### 3.2.4 Prevalence of malaria and anaemia according to the use of preventive measures

As part of their routine ANC visits, pregnant women are given insecticide treated nets (ITNs) during registration and monthly Sulphadoxine-pyrimethamine (SP) as intermittent preventive treatment (IPT).

Sixty-two (27.9%) participants admitted to having taken SP during ANC visit. IPT-SP users had higher prevalence in both malaria (25.8%) and anaemia (33.9%) compared with non-IPT users (17.5% and 24.4% respectively). However, there was no statistically significant difference between the two cases (p = 0.165 and p = 0.153 respectively) (Table 3). IPT usage was higher among multigravidae than the primigravidae. Participants in the third trimester recorded the highest for IPT usage and those in the first trimester recording the least. There was a significant difference between the three groups (p<0.0001). Table 2 shows the usage of IPT among participants with respect to gravidity and term of pregnancy.

One hundred and forty-six (65.8%) participants possessed an ITN with 72 (49.3%) of them sleeping under it the previous night (ITN users). Malaria prevalence was higher (20.5%) amongst ITN owners whilst that of the non-owners was 18.4%. Thirty-three (22.6%) of ITN owners were anaemic compared to twenty-seven (35.5%) of non-owners; and the difference was significant (p=0.040). Among ITN users, malaria prevalence was 19.4% and anaemia prevalence 20.8%. For ITN non users, prevalence was 20.0% and 30.0% for malaria and anaemia respectively (Table 3).

### 3.3 Follow up group

One hundred participants were recruited for the longitudinal cohort study (follow up). Due to reasons such as migration, change of ANC visit dates and opting out by some participants for personal reasons, fifty-six participants were left for the follow up study. Unfortunately, two of the fifty-six also experienced spontaneous abortion. A total of fifty-four participants were therefore successfully followed up until delivery.

With one hundred and thirty-nine (139) tests performed, the overall prevalence of malaria and anaemia among the participants involved were 18.7% and 32.4% respectively. At delivery, twenty-two (40.7%) out of these, had malaria parasitaemia at least once during the pregnancy period. Twenty-eight (51.9%) of them were also anaemic at least once during this period. There were twenty-six episodes of malaria among participants who were followed up till delivery. For anaemia, there were forty-five episodes of which twenty-one (46.7%) were malaria associated. Women who had malaria during pregnancy had increased risk of anaemia (OR=10.20 95% CI= 2.840-36.630, p <0.001) (Table 4).

**Table 4:** Maternal characteristics of participants involved in follow up

|  | Participants | Malaria |  | Anaemia |  | LBW | Mean Birthweight |  |
| --- | --- | --- | --- | --- | --- | --- | --- | --- |
|  | n (%) | n (%) | p-value | n (%) | p-value | n (%) | Kg±SD | p-value |
| **Total** | 54 (100) | 22 (40.7) |  | 28 (51.9) |  | 3 (5.6) | 3.185 ±0.5461 |  |
| **Age** |  |  |  |  |  |  |  |  |
| <20 | 4 (7.4) | 2 (50.0) | 0.695 | 2 (50.0) | 0.375 |  | 2.950 ±0.3697 | 0.296 |
| ≥20 | 50 (92.6) | 20 (40.0) |  | 26 (52.0) |  | 3 (6.0) | 3.204 ±0.5562 |  |
| **Gravidity** |  |  |  |  |  |  |  |  |
| Primigravida | 15 (27.8) | 8 (53.3) | 0.243 | 8 (53.3) | 0.939 |  | 3.133 ±0.3697 | 0.669 |
| Multigravida | 39 (72.2) | 14 (35.9) |  | 20 (51.3) |  | 3 (7.7) | 3.205 ±0.6035 |  |
| **ANC visits** |  |  |  |  |  |  |  |  |
| <4 | 2 (3.7) | 2 (100) | 0.082 | 1 (50.0) | 0.957 |  | 3.050 ±0.7778 | 0.805 |
| ≥4 | 52 (96.3) | 20 (38.5) |  | 27 (51.9) |  | 3 (5.8) | 3.190 ±0.4289 |  |
| **IPT dosage** |  |  |  |  |  |  |  |  |
| <3 | 24 (44.4) | 11 (45.8) | 0.496 | 14 (58.3) | 0.394 | 2 (6.7) | 3.275 ±0.6635 | 0.364 |
| ≥3 | 30 (55.6) | 11 (36.7) |  | 14 (46.7) |  | 1 (4.2) | 3.113 ±0.4289 |  |
| **Malaria** |  |  |  |  |  |  |  |  |
| Positive | 22 (40.7) |  |  | 18 (81.8) | < 0.001 | 2 (9.1) | 3.259 ±0.7076 | 0.415 |
| Negative | 32 (59.3) |  |  | 10 (31.3) |  | 1 (3.1) | 3.134 ±0.4053 |  |
| **Anaemia** |  |  |  |  |  |  |  |  |
| Anaemic |  |  |  |  |  | 3 (10.7) | 3.250 ±0.6546 | 0.9708* |
| Non-anaemic |  |  |  |  |  |  | 3.115 ±0.3997 |  |
| Malaria-associated anaemia |  |  |  |  |  | 2 (11.8) | 3.282 ±0.7683 |  |

### 3.4 Birthweights of babies

All deliveries were singleton with birthweight ranging from 1.9 to 5.4 kg. The mean birthweight of babies of all participants was 3.185 kg (±0.5461). Three (5.6%) babies out of 54 had low birthweight (<2.5 kg) and there was one case (1.9%) of macrosomia (birthweight ≥ 4.0 kg). Table 4 shows maternal characteristics and their effects on birthweight in the follow up group.

#### 3.4.1 Effects of malaria and anaemia on birthweight

The mean of birthweights was higher in participants who were parasitaemic (3.259 kg ±0.7076) and anaemic (3.250 kg ±0.6546) during the pregnancy period (Table 4). However, there was no significant difference in mean birthweights between parasitaemic and non-parasitaemic women. The same was observed for anaemic and non-anaemic women. The mean birthweight of babies born to mothers who had malaria-associated anaemia was 3.282 kg ±0.7683 and there was no significant difference between them and that of babies born to non-malaria-associated anaemic mothers (Table 4).

##### 3.4.1.1 Effects of frequency of malaria and anaemia on birthweight

Out of the total (22) who had malaria during the pregnancy period, 3 had the infection more than once. In comparing the means of birthweight, participants who had malaria once during the period had a higher mean birthweight (3.217 kg ±0.4878) than those who had the infection more than once (2.800 kg ±0.7810). The same trend was seen for anaemia where participants who were anaemic once (14 (50%)) in the pregnancy period had a higher mean birthweight (3.179 kg ±0.4191) than those who were anaemic more than once (3.162 ±0.6117) (Table 5). Nonetheless, there were no significant difference in mean birthweights between mothers in both malaria and anaemia. Participants who had malaria-associated anaemia once during the pregnancy period also had a higher mean birthweight (3.231 kg ±5006) compared to those who had it twice or more (2.800 kg ±7810). This difference was also not statistically significant.

**Table 5:**
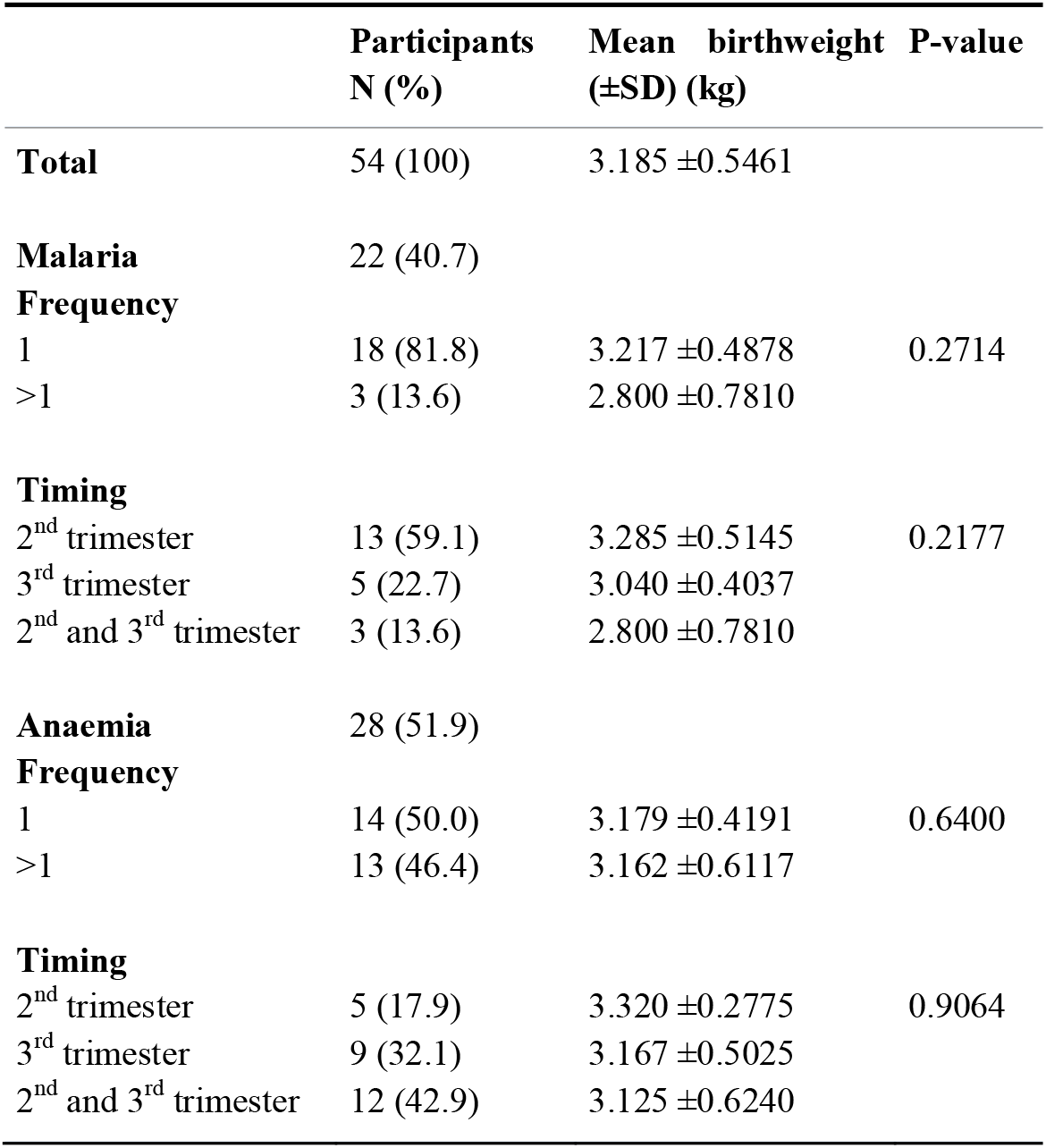
Effects of frequency and timing of malaria and anaemia on birthweight of babies

| | Participants<br>N (%) | Mean birthweight<br>(±SD) (kg) | P-value |
| --- | --- | --- | --- |
| **Total** | 54 (100) | 3.185 ±0.5461 | |
| **Malaria** | 22 (40.7) |  |  |
| **Frequency** |  |  |  |
| 1 | 18 (81.8) | 3.217 ±0.4878 | 0.2714 |
| >1 | 3 (13.6) | 2.800 ±0.7810 | |
| **Timing** |  |  |  |
| 2<sup>nd</sup> trimester | 13 (59.1) | 3.285 ±0.5145 | 0.2177 |
| 3<sup>rd</sup> trimester | 5 (22.7) | 3.040 ±0.4037 | |
| 2<sup>nd</sup> and 3<sup>rd</sup> trimester | 3 (13.6) | 2.800 ±0.7810 | |
| **Anaemia** | 28 (51.9) |  |  |
| **Frequency** |  |  |  |
| 1 | 14 (50.0) | 3.179 ±0.4191 | 0.6400 |
| >1 | 13 (46.4) | 3.162 ±0.6117 | |
| **Timing** |  |  |  |
| 2<sup>nd</sup> trimester | 5 (17.9) | 3.320 ±0.2775 | 0.9064 |
| 3<sup>rd</sup> trimester | 9 (32.1) | 3.167 ±0.5025 | |
| 2<sup>nd</sup> and 3<sup>rd</sup> trimester | 12 (42.9) | 3.125 ±0.6240 | |

##### 3.4.1.2 Effects of timing of malaria and anaemia on birthweight

The timing of the infection and anaemia in terms of trimester in which they occurred were also taken. For malaria, 13 (59.1%) of infection occurred in the second trimester compared to 5 (22.7%) in the third trimester. Three participants had malaria in both the second and third trimesters accounting for 13.6% of total infections. Amongst participants who had anaemia during the pregnancy period, the highest number (42.9%) of them were anaemic in both the second and third trimesters compared to 17.9% and 32.1% in the second and third trimesters respectively (Table 5).

The participants who had the infection in the second trimester alone had the highest mean birthweight of babies (3.285 kg ±0.5145) followed by third trimester alone (3.040 kg ±0.4037) and both second and third trimesters (2.800 kg ±0.7810). The same trend was seen for anaemia. However, these were not statistically significant (Table 5).

## 4.0 Discussion

Close to half of the participants involved in the present study did not take the recommended dose of SP during the pregnancy period, especially those from the rural areas. This could be due to the lifestyle of the pregnant women attending ANC as and when they feel ill instead of the routine planned dates of visits. This in turn exposes the pregnant women and their unborn babies to the adversities of malaria.

Being pregnant alone is a major factor that compromises the immune response of women, increasing their vulnerability to majority of infectious diseases (Sappenfield et al., 2013; Tay et al., 2017). Therefore, during pregnancy, the infection poses a high risk to the mother especially the primigravidae (Valente et al., 2011). Risks associated with malaria during pregnancy include foetal death, premature delivery, intra-uterine growth retardation, low birthweight, maternal anaemia and mortality (Wini et al., 2013).

The overall prevalence of malaria at enrolment was 19.8%. Ofori et al., (2009) and Enato et al., (2009) recorded similar prevalence amongst pregnant women in the Greater Accra region and Nigeria respectively. This is higher than the prevalence of the infection that was recorded in several studies in Ghana among comparatively larger sample sizes (Ampofo et al., 2018; Helegbe et al., 2018; Osarfo et al., 2017). Other studies in Angola and Malawi have respectively reported 8.6% (Valente et al., 2011) and 11.9% (Rogawski et al., 2012) of the infection among pregnant women. The higher prevalence recorded in the present study could be due to participants who came from the rural areas as hospitals that included more of such participants had increased odds of infection. However, there are other studies that have reported higher prevalence than what has been reported in this study (Agan et al., 2010a; Akinboro et al., 2010; Anchang-Kimbi et al., 2015). This is because these studies were conducted in stable transmission areas, majority of pregnant women coming for the first time and also registering late at the ANC.

Van-Spronsen *et al*., (2012) found a prevalence of anaemia (27%) similar to that of the present study. A relatively higher pregnancy-associated anaemia has been reported in earlier studies in the country, in the Greater Accra region (Tay et al., 2017) and in Nigeria (Agan et al., 2010b) with a 66.4% and 59.6% prevalence respectively. Malaria-associated anaemia was 54.5% among participants in the present study. Also, malaria-positive women were more likely to have anaemia and this association was statistically significant with a p-value <0.001. However, not all pregnant women who were anaemic were malaria positive. This could be as a result of other factors that cause anaemia during pregnancy such as HIV, helminth infections, folate and nutritional deficiencies (Glover-Amengor et al., 2005).

Agona and Kuntanase Government hospitals are in peri-urban settings and have most of their attendants coming from neighbouring rural communities. Malaria transmission is affected by location and rural populations are at greater risk of the infection (Rogerson et al., 2018). This is due to the availability of suitable environments for vector breeding from poor sanitation and the absence of appropriate control measures (Kimbi et al., 2013; Molina Gómez et al., 2017). Thus, the higher infection prevalence recorded in these hospitals than in KNUST hospital and Aniniwah Medical Centre. However, it must be stated that even though KNUST hospital and Aniniwah Medical Centre are both located in an urban area, approximately 6.4 km apart, the prevalence of malaria and anaemia recorded in the latter is far greater than the former. This could be attributed to difference in several factors including lifestyle of pregnant women who reported to both hospitals which may increase or decrease their vulnerability to both malaria and anaemia. Moreover, majority of the pregnant women reporting to both hospitals were attending ANC for the first time. Future studies can look into factors that may be responsible for these differences.

In this study, it was observed that younger pregnant women were more prone to both malaria and anaemia than their older counterparts with higher prevalence. There was a significant correlation between both malaria (p=0.027) and anaemia (p=0.026) in younger and older pregnant women. Also, primigravid women were found to be more parasitaemic compared to their multigravida counterparts. A study in Nigeria also reported similar prevalence of malaria (24.3%) among primigravid women (Enato et al., 2009). However, the difference in the present study was not significant. This is in disagreement with findings of a study which found a significantly higher parasitaemia in primigravid women (Agan et al., 2010a). Most of the younger pregnant women (95.2%) in this study were primigravidae. Being young and primigravid have been reported to be a risk factor for malaria infection and its associated anaemia in several studies (Anchang-Kimbi et al., 2015; McClure et al., 2014; Orish et al., 2012; Tay et al., 2013). Compared with their multigravida counterparts, primigravid women are also known to be vulnerable to infections especially malaria (Rogerson et al., 2018). Due to multiple pregnancies, multigravida women tend to be exposed to the infection for longer periods with occasional infection of their placentas, therefore develop partial immunity in successive pregnancies (Moya-Alvarez et al., 2014; Wassmer and Grau, 2016). Furthermore, experience and knowledge of malaria and its prevention during pregnancy by multigravida woman could also be an advantage. Although the level of immunity is a determinant of the susceptibility to, and severity of malaria, it is also affected by the infection intensity and transmission stability (Sappenfield et al., 2013).

With gestational age, women in the second trimester had the highest prevalence of the infection. Similar observation was made by Afrifa et al., (2017) amongst malaria positive pregnant women in Koforidua, in the Eastern region of Ghana. It is recommended by WHO to administer IPT-SP in the 16^th^ week of pregnancy, which is in the second trimester (Wini et al., 2013). Therefore, under normal circumstances, this is the stage at which treatment of malaria infection would take place thereby reducing the infection prevalence. Some studies also found that pregnant women have increased chances of getting infected in the first half of pregnancy which is within the first and second trimesters (Huynh et al., 2011; Valea et al., 2012).

The prevalence of anaemia was highest in participants in their third trimester which is similar to findings by Tay et al., (2013). In the third trimester of pregnancy, anaemia has been reported to occur in about one third of women with major causes being iron and folate deficiencies (Friel, 2020). On the contrary, other studies reported reduced prevalence of anaemia in the third trimester (Anlaakuu and Anto, 2017; Nega et al., 2015). Prevalence of anaemia has been reported to be different in different countries and also different within regions of a country (Koyuncu et al., 2017). Difference in national policies and socio-economic status may be accountable for this observation.

The World Health Organization recommends intermittent preventive treatment with sulphadoxine-pyrimethamine (IPT-SP) together with the use of insecticide-treated nets as a means of protection of women from malaria during pregnancy (WHO, 2014).

The national coverage target of IPT-SP and ITN is 80% (Orish et al., 2015; Stephen et al., 2016). In the present study, IPT-SP coverage was 27.9% at enrolment and this included participants who were coming to ANC for the first time as well as participants who were in the early stage of pregnancy (below 16 weeks). Comparing IPT-SP users and non-users, IPT-SP users had higher prevalence of both malaria and anaemia. This could be as a result of exposure of IPT-SP users to repeated infectious mosquito bites thereby resulting in infection and also other factors which may lead to anaemia such as helminth infection, iron and folate deficiencies. Nonetheless, this difference was not statistically significant. Orish et al., (2015) also found IPT-SP users to have higher prevalence of anaemia compared to non-users. IPT-SP is known to be effective in reducing maternal parasitaemia and its associated complications such as anaemia (Esu et al., 2018). Other studies found that IPT users had less parasitaemia and anaemia compared to IPT non-users (Tutu et al., 2010; Wilson et al., 2011).

As part of strategies to eliminate malaria, protective measures such as protection from mosquito bites is an integral part. As such, in Ghana, as part of ANC registration, insecticide treated nets are given in efforts to protect the pregnant woman and her unborn child. In the present study, at enrolment, ITN ownership was 65.8% but usage was 49.3% the night before their recruitment into the study. Osarfo et al., (2017) also reported 60% ITN ownership and 40% ITN usage. Reasons for not owning or using the nets included hot conditions inside the net and the use of other vector control measures. Similar complaints were made by pregnant women in a study in the Brong Ahafo region of Ghana (Manu et al., 2017).

The prevalence of malaria amongst ITN owners was 20.5% and in those who slept under it the night before, 19.4%. Amongst non-owners, the prevalence (18.4%) was lower. These differences, however, were not statistically significant. Thus, using ITN did not provide extra protection to the pregnant women. This could be attributed to infectious vector bites prior to the use of the net or even after use, as a result of different lifestyle behaviours or in cases where there are no other control measures used. Similar observations were made by Browne et al., (2001).

In the longitudinal study, there was a follow-up on fifty-four pregnant women up to delivery of their babies. The mean number of ANC visits was 8.26 ± 2.665 which was higher than what was observed in other studies in the country (Asundep et al., 2014; Völker et al., 2017). Antenatal Care (ANC) is an integral tool in the improvement of maternal and neonatal health through the early detection and management of complications during pregnancy (Sumankuuro et al., 2017). In the present study, 52 (96.3%) visited ANC four times or more which is the least recommendation for uncomplicated pregnancy (Asundep et al., 2014). This was higher than the findings of a study which reported 74.6% of participants attending ANC four or more times (Addai-Mensah et al., 2018). Malaria was more prevalent among this group of pregnant women and this could be as a result of the reduced number of SP taken as none was able to take the recommended dosage. However, there was no significant difference between the two groups. Increasing ANC visits have been reported to provide the opportunity for increased uptake of SP thereby increasing protection (Nkoka et al., 2018).

IPT–SP uptake among participants in the follow up study was 100% with only 55.6% (30) out of this taking the recommended 3 or more dosages. This, however, did not have any significant impact on the prevalence of malaria and anaemia in both groups. Increase in SP taken during pregnancy is reported to minimize malaria and its associated complications such as anaemia and low birthweight (Valea et al., 2012). A single dose of SP has been reported to provide protection from malaria and its consequences. This is supported by findings from the present study and that of Tutu et al., (2011b).

In Sub-Saharan Africa, malaria and anaemia are major contributors to poor birth outcomes such as spontaneous abortion, preterm delivery and low birthweight (Tutu et al., 2011a). In the present study, the poor birth outcomes recorded were spontaneous abortion and low birthweight. The overall mean birthweight for all babies in the present study was 3.185 kg (±0.5461). This is higher than that reported from Benin (2.9982 kg ±0.474) (Huynh et al., 2011). Low birthweight was seen in three (5.6%) of the babies, a mean birthweight of 2.067 kg (±0.1528). Previous studies have reported higher percentages of low birthweight babies among maternal cohorts in Ghana (Laar et al., 2013) and in Benin (Huynh et al., 2011). However, another study conducted in the Greater Accra region of Ghana recorded a lower LBW prevalence of 3.3% (Stephens et al., 2014).

In the present study, it was observed that neither time of infection nor its frequency had a significant impact on the mean birth weight of babies. However, Huynh et al., (2011) reported that mothers who had an infection in the second trimester had higher mean birthweight babies compared to those who had infection in the third trimester. Similar observation was also made regarding whether or not a mother took the recommended dose of SP as it had no impact on the mean birthweight of babies. This is in disagreement with a study in Cameroon which found a higher mean birthweight of babies born to pregnant women who took the full course of IPT (3 doses) (Yoah et al., 2018). Increasing dosage of IPT-SP has been found to be associated with higher mean birthweight and reduces the incidence of LBW (WHO, 2014). However, according to Tutu et al., (2011b), taking a single dose of SP can improve the birthweight of babies.

Overall, in the present study, malaria and anaemia had no significant impact on birthweight of babies. Similar observation was made by another study which found no association between anaemia and low birthweight (Port et al., 2012). This could be attributed to the lower number of low birthweights recorded. Furthermore, the lower number of low birthweight babies and the less impact of both malaria and anaemia could be attributed to the 100% uptake of SP and ITN usage among the follow-up pregnant women during their pregnancy period. These have been reported to provide extra protection to women during pregnancy. Also, a higher percentage of women visited ANC for the recommended number for uncomplicated pregnancy.

Nonetheless, several studies have found malaria and anaemia to be significantly associated with low birthweight (Guyatt and Snow, 2004; Nair et al., 2018; Wini et al., 2013).

## 5.0 Conclusion

The findings from this study suggest that malaria and anaemia are still prevalent among pregnant women. Malaria was significantly associated with anaemia during pregnancy. Pregnant women of age lesser than 20 years as well as rural dwellers are at higher risk of malaria. Low birth weight was lower among babies born to the pregnant women. Malaria and anaemia did not have a significant impact on the birthweight of the babies. ITN and IPT-SP usage were also high but had no significant impact on malaria, anaemia and birthweight of babies born to the pregnant women. Education of women of child bearing age on the possible risk of malaria and anaemia during pregnancy on the pregnant woman and her foetus, should be intensified. Furthermore, education on malaria prevention and use of preventive measures especially in rural areas should be encouraged.

## Data Availability

All the underlying data for this manuscript has been deposited in Open Science Framework https://osf.io/cf9jw/. Scientist can use without restrictions provided they give the appropriate credits and citations. The citation of this data can be accessed from the DOI: link here https://doi.org/10.17605/OSF.IO/CF9JW
Badu, K. (2021, August 11). Plasmodium falciparum malaria during pregnancy: the impact of parasitaemia and anaemia on birthweight. https://doi.org/10.17605/OSF.IO/CF9JW

## Authors’ contributions

**Dawood Ackom Abbas**: Conceptualization, Methodology, Resources, Investigation, Formal analysis, writing – original draft; **Kingsley Badu and Bernard Walter Lartekwei Lawson**: Conceptualization, Methodology, Resources, Validation, Supervision, Writing - Review & Editing; **Abdul-Hakim Mutala, Samuel Kekeli Agordzo and Christian Kwasi Owusu**: Investigation, Formal analysis, Writing - Review & Editing

## Acknowledgements

The authors would like to thank all pregnant women who participated in the study. Special gratitude to midwives and laboratory technicians in the respective study areas.

